# Circulating Microparticles are increased amongst people presenting with HIV and advanced immune suppression in Malawi and correlate closely with arterial stiffness

**DOI:** 10.1101/2020.07.19.20157107

**Authors:** C Kelly, R Gurung, W Tinago, R Kamngona, I Sheha, M Chammudzi, K Jambo, J Mallewa, A Rapala, RS Heyderman, PWG Mallon, H Mwandumba, S Khoo, N Klein

## Abstract

**Objectives:** We aim to investigate whether circulating microparticle (CMPs) subsets were raised amongst people presenting with HIV and advanced immune suppression in Malawi, and whether they associated with arterial stiffness.

**Methods:** ART-naïve adults with a new HIV diagnosis and CD4 <100 cells/µL had microparticle characterisation and carotid femoral Pulse Wave Velocity (cfPWV) at 2 weeks post ART initiation. HIV uninfected controls were matched on age, systolic BP and diastolic BP in a 1:1 ratio. Circulating microparticles were identified from platelet poor plasma and stained for endothelial, leucocyte, monocyte and platelet markers.

**Results:** The median (IQ) total CMP count was 1 log higher in participants with HIV compared to those without (p<0.0001) and was associated with arterial stiffness (spearman rho 0.47, p<0.001). In adjusted analysis, every log increase in circulating particles showed a 20% increase in cfPWV (95% CI 4 – 40%, p=0.02).

In terms of subsets, endothelial and platelet derived microparticles were most strongly associated with HIV. Endothelial derived E-selectin+ CMPs were 1.3log-fold higher and platelet derived CD42a+ CMPs were 1.4log-fold higher (both p<0.0001). Endothelial and platelet derived CMPs also correlated most closely with arterial stiffness [spearman rho: E-selectin+ 0.57 and CD42a 0.56, both p<0.0001).

**Conclusions:** Circulating microparticles associate strongly with arterial stiffness among PLWH in Malawi. Endothelial damage and platelet activation are of particular importance and future translational studies should consider prioritising this pathway.

## Introduction

People living with HIV (PLWH) are at increased risk of cardiovascular diseases (1–4). Heightened inflammation has been implicated in vascular dysfunction but the underlying pathogenetic mechanisms have not been fully elucidated (2). These mechanisms are even more complex in the low-income sub-Saharan Africa setting (SSA), where additional factors including latent or recurrent infections, late presentation and ART failure are prevalent (5–7). In Malawi, several studies have shown an increased risk of stroke associated with HIV (8, 9).

The first few months of antiretroviral treatment (ART) carry a particularly high cardiovascular risk. In a case control study of 705 patients with stroke, Benjamin *et al* found an adjusted odds ratio of 15.6 among those within the first 6 months of ART compared to those not yet started on treatment and 12.9 among those with a CD4 count of less than 200 cells/mm^3^ compared to those with CD4 greater than 500 cells/mm^3^ (9). Further, we have previously shown that amongst people presenting with HIV and a CD4 count less than 100 cells/mm^3^, arterial stiffness is Increased by 12% (adjusted fold change) compared to healthy volunteers living without HIV (5). Arterial stiffness has been used as a measure of vascular dysfunction in HIV studies as well as other chronic inflammatory conditions (10, 11) and we have also previously shown that arterial stiffness, as measured by carotid femoral pulse wave velocity (cfPWV), is increased in people experiencing unstructured treatment interruption (12). cfPWV is a gold standard measurement of arterial stiffness and has been validated as a physiological biomarker of cardiovascular events and mortality (13, 14).

The increase in arterial stiffness previously Identified only persists during the first 3 months of ART and is associated with markers of inflammation, including T cell activation. Given the timing of this inflammatory process, it is tempting to hypothesise that increased cardiovascular risk during this time may follow a similar process to Immune Reconstitution Inflammatory Syndrome (IRIS). During the normal inflammatory response leucocytes adhere to the endothelial cells via VCAM, ICAM, and undergo transcytosis leading to the development of foam cells (15). Pro-thrombotic pathways attract platelets to the resulting plaque and subsequent migration of smooth muscle cells into the intima leads to the formation of a fibrous cap (16). During this process, elastin is degraded by enzymes such as MMPs and is replaced by collagen, increasing arterial stiffness (16).

Circulating Microparticles (CMPs) are released into the circulation following activation or apoptosis of the affected cells (17). Through a process of blebbing, microparticles are formed from the originating host cell’s outer membrane (e.g. leucocytes, platelets and endothelial cells). During this process annexin V molecules, which are normally located on the inner membrane of a cell, are flipped round to become expressed on the microparticle surface (18). These microparticles also express the markers expressed on the surface of the cell of origin and so microparticle subsets are an indication of which cells are undergoing stress.

Previous studies in HIV have assessed the presence and function of CMPs amongst PLWH in high resource settings and have separately highlighted tissue factor expression, imbalanced endothelial progenitor cell proportions and platelet activation (19–22). Hijmans *et al* recently demonstrated evidence of in vitro endothelial cell stress, apoptosis and senescence induced by leucocyte, platelet and endothelial microparticle subsets from patients with HIV (23). Here, we aim to take the characterisation of circulating microparticles in HIV a step further by assessing their relationship with cardiovascular risk and seeking to elucidate pathways that might be involved in heightened inflammation during early ART.

## Methods

### Study cohort

The Study into HIV, Immune activation and EndotheLial Dysfunction (SHIELD) cohort recruited ART-naïve adults with a new HIV diagnosis and CD4 <100 cells/µL from the ART clinic and voluntary HIV testing clinic at Queen Elizabeth Central Hospital, Blantyre, Malawi, along with adults without HIV infection and without evidence of infection within the previous 3 months. This cohort has been reported on previously, and full details of recruitment are available in (5). In brief, participants underwent a detailed clinical assessment, blood draw for plasma storage and cfPWV at 2 weeks post ART initiation. cfPWV measured arterial stiffness using a Vicorder device (Skidmore Medical, London, UK) according to standardised guidelines (24). A random sample of wave forms was reviewed by an experienced independent assessor at three timepoints during the study to ensure consistent quality.

All participants provided informed written consent and ethical approval was granted by the College of Medicine Research and Ethics Committee (COMREC), University of Malawi (P.09/13/1464) and the University of Liverpool Research and Ethics Committee (UoL000996).

SHIELD participants with and without HIV infection were ordered according to cfPWV values from lowest to highest. A convenience sample of 36 PLWH and 36 HIV matched controls without HIV infection were chosen from across the spectrum of cfPWV values. Participants with a cfPWV in the highest quartile (>8.2 m/s) were chosen randomly in a 2:1 ratio to patients with a cfPWV below the highest quartile from the cohort of patients with HIV. This was to enrich the number of potential CMPs for subset analysis (we hypothesised they would be higher at higher ranges of cfPWV) whilst also capturing a range of values to analyse associations between total CMP count and cfPWV. Participants without HIV were then matched to participants with HIV infection on age, systolic BP and diastolic BP in a 1:1 ratio.

### Statistical analysis

Wilcoxon Ranksum and spearman rho were used to test association between CMPs and categorical or continuous variables respectively. To allow for multiple comparisons characterising 18 types of CMPs, a p value of less than 0.003 was used as significant. Linear regression was used to examine the effect of total CMPs on cfPWV which was log transformed for normality. The model was adjusted for confounders (age and sex) and mediators (blood pressure, haemoglobin and HIV) as identified previously (5).

### Laboratory procedures

Platelet poor plasma (PPP) was isolated from thawed plasma and stained for Annexin V (supplementary material). Endothelial, leucocyte, monocyte and platelet microparticles were chosen as common circulating microparticles involved in inflammation and were identified by flow cytometry on a CyAn ADP 9 colour flow cytometer (Beckman Coulter). FITC stained for Annexin V (BD Pharmingen), PE stained for VCAM, ICAM, E-selectin, 66b or CD16 (BD Pharmingen) and APC-Cy7 stained for PECAM or CD14 (BD Pharmingen). To identify tissue factor expression on monocytes, PE stained Annexin V, APC-Cy7 stained CD14 (BD Pharmingen) and FITC stained tissue factor (Sekisui Diagnostics). Single stain samples were also acquired for the purposes of compensation and isotype controls were analysed for gating. After identification of singlets, the microparticle pellet was gated on forward scatter and AnV (FITC) to identify the microparticle population which was less than 1µm in size and expressing AnV (see Figure 1 **Error! Reference source not found.**). Gates were applied using thresholds provided by isotype controls.

**Figure 1.**
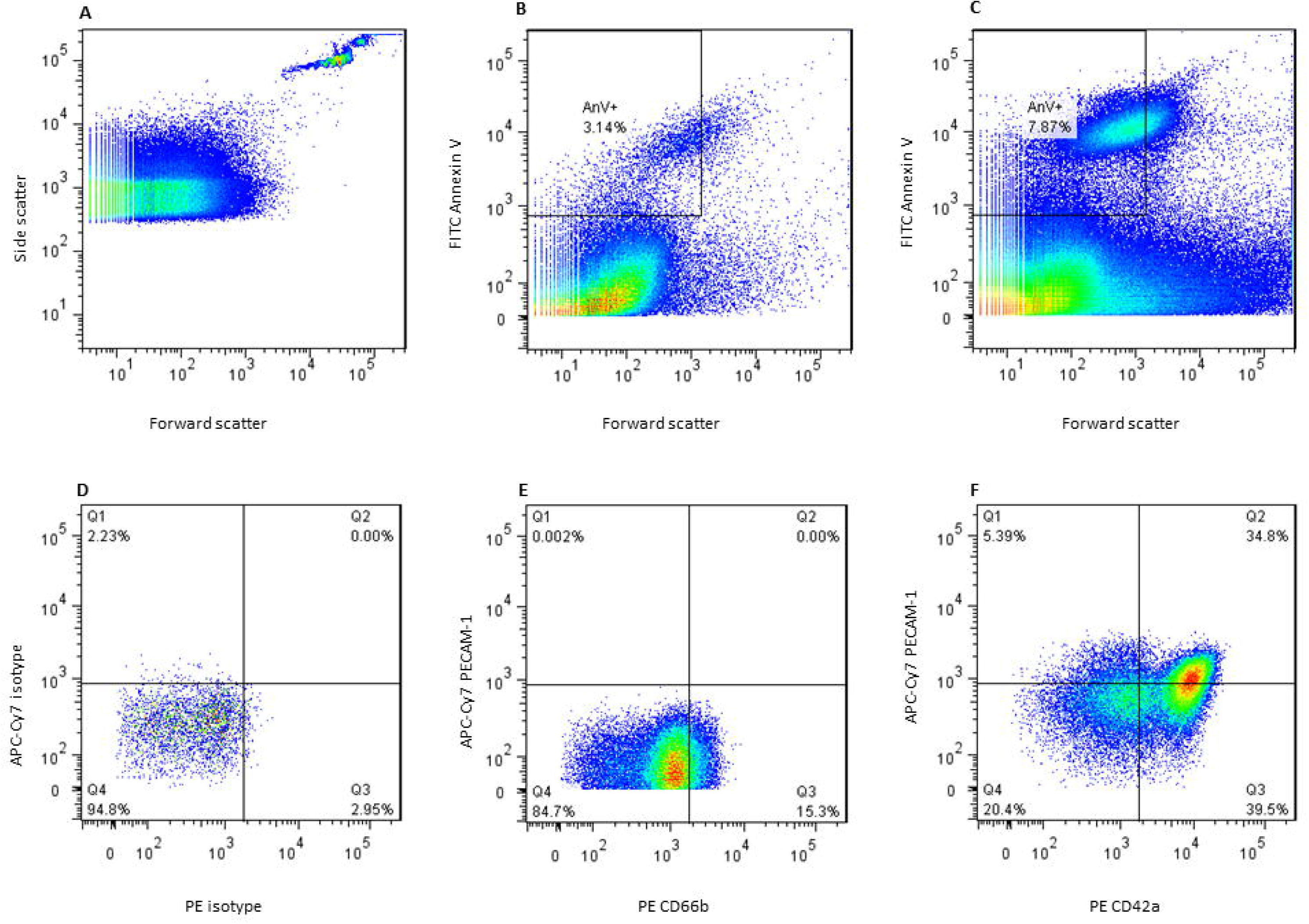
Identification of the microparticle population and subset gating strategy. **Figure 1 legend. A**. 1.1µm beads were used to find forward scatter gate for microparticle identification **B**. Microparticle pellet was obtained from platelet poor plasma and stained for Annexin V. Total circulating microparticle count was measured by identifying microparticles Annexin V positive and less than 1.1µm forward scatter as shown. **C**. Total circulating microparticles in a patient with a high count compared to **B. D**. Isotope controls identified gates for PE and APC-Cy7 stains for each subset panel. E. Leucocyte microparticles F. Endothelial or platelet microparticles (see table 4).

Cell surface immune phenotyping was carried out for CD4 and CD8 T cell activation (HLA-DR+/CD38), exhaustion (PD1+) and senescence (CD57+), as well as monocyte subsets (classical CD14 CD16, intermediate CD14 CD16 and nonclassical CD14 CD16). The T-cell panel consisted of CD3 BV510, CD4 V450, CD38 PE Cy7, HLA-DR AF700, PD1 APC, and CD57 FITC (all from BD Biosciences) and CD8 PE (Biolegend). The monocyte panel consisted of HLA-DR AF700, CD14 PE Cy7 and CD16 PE (all from BD Biosciences).

## Results

CMP data were available for 33 PLWH and 36 people without HIV. Sufficient plasma samples were not available for three of the selected participants with HIV. For the 69 participants with available CMP data, median (IQR) age was 41 years (35 – 49) and 28 (41%) were female. 31 (45%) had an education level of at least primary school completion. Median (IQR) blood pressure was 130/78 (118/70 – 134/80) and 11 (16%) had a history of smoking. Characteristics according to HIV status are shown in Table 1 (for in depth comparison according to HIV status see (5)). For the 33 PLWH, median (IQR) CD4 and HIV viral load were 42 cells/µL (31 – 71) and 1.1×10 copies/mL (0.4 – 2.6).

**Table 1.**
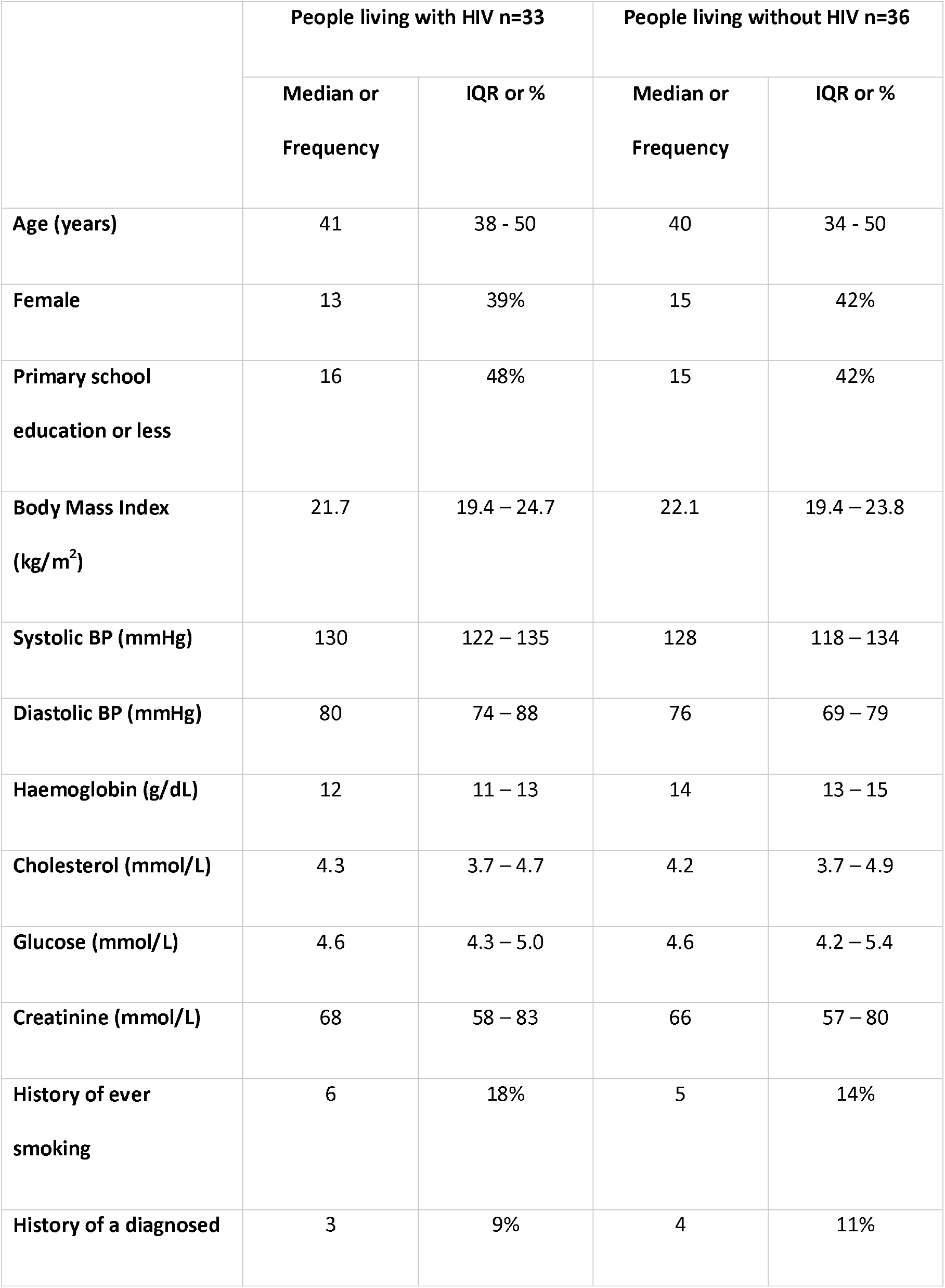
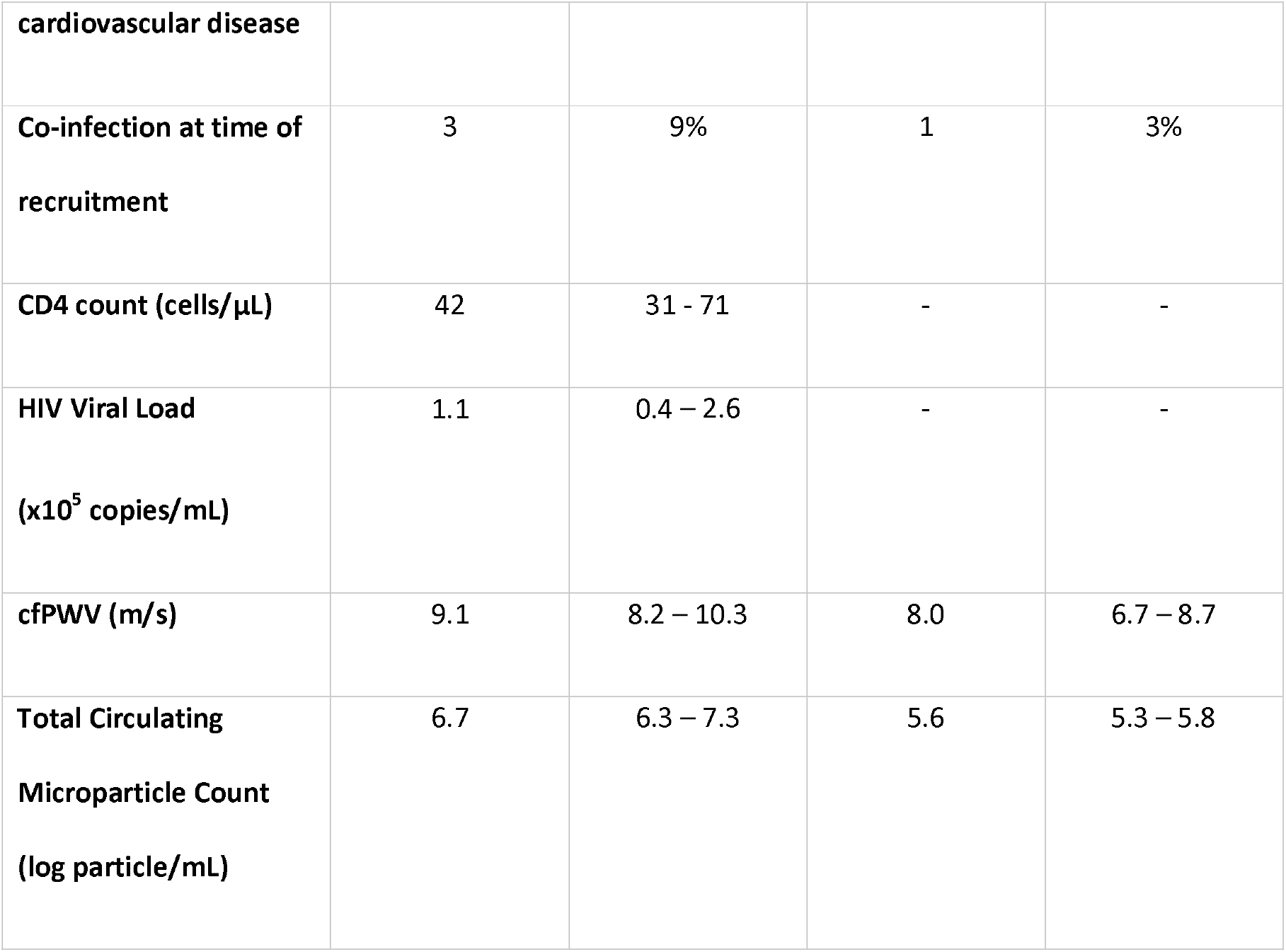
Clinical characteristics of 67 SHIELD participants with microparticle data according to HIV status.

### Relationship between absolute CMP counts, HIV and arterial stiffness

For the overall cohort, the median (IQ) total CMP count was approximately 1 log higher in participants with HIV compared to those without (table 1). Total CMP counts were also significantly associated with arterial stiffness (spearman rho 0.47, p<0.001; figure 2), as well as faster heart rate and higher creatinine (spearman rho: 0.31; p=0.01 and 0.28; p=0.02). When adjusted for *a priori* identified mediators and confounders (age, sex, haemoglobin and blood pressure), higher log total CMPs predicted increased cfPWV [fold change 1.20m/s, 95% CI 1.04 – 1.40; p=0.02]. When additionally adjusted for HIV status, the effect of log total circulating microparticle count on cfPWV remained similar [fold change 1.07m/s (95% CI 1.23 – 1.01), p=0.046. Table 2].

**Figure 2.**
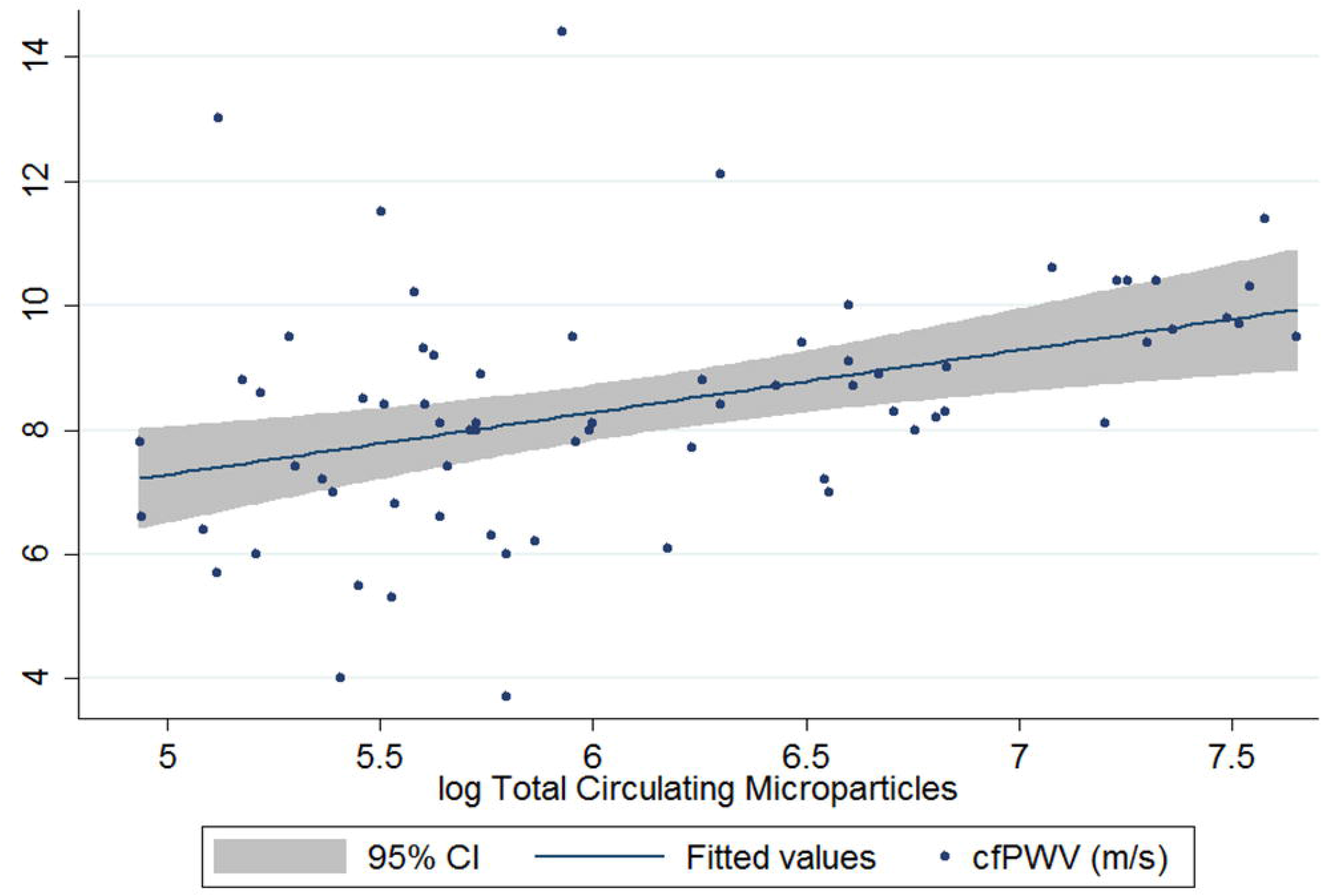
Association between Total Circulating Microparticles and carotid femoral Pulse Wave Velocity (m/s)

**Table 2.** Linear regression analysis showing the effect of total circulating microparticles on carotid femoral Pulse Wave Velocity **Table 2 Legend:** A multivariate model was constructed to assess the effect of circulating microparticles (CMPs) on carotid femoral Pulse Wave Velocity (cfPWV) using data from patients with and without HIV combined. The models are adjusted for mediators and confounders as identified from previous work in this cohort (5). The first model provides the effect of CMPs on arterial stiffness adjusting for mediators and confounders but excluding HIV. The second model adds HIV to assess for any role of HIV in mediating the effect of CMPs on arterial stiffness.

|  | Effect of total circulating microparticle count<br>on cfPWV |  |  | Effect of total circulating microparticle count on cfPWV<br>adjusting for HIV |  |  |
| --- | --- | --- | --- | --- | --- | --- |
|  | Fold change | 95% CI | P value | Fold change | 95% CI | P value |
| **Total Microparticle Count**<br>(per log particle/mL increase) | 1.20 | 1.04 – 1.40 | 0.02 | 1.23 | 1.01 – 1.50 | 0.046 |
| **Age**<br>(per 10 year increase) | 1.14 | 1.03 – 1.26 | 0.02 | 1.12 | 1.02 - 1.24 | 0.03 |
| **Female sex** | 0.85 | 0.67 – 1.07 | 0.18 | 0.84 | 0.66 – 1.07 | 0.17 |
| **Diastolic Blood pressure (per 10mmHg increase)** | 1.20 | 1.06 – 1.38 | 0.004 | 1.21 | 1.07 – 1.38 | 0.004 |
| **Haemoglobin**<br>(per g/dL increase) | 1.04 | 0.98 – 1.10 | 0.19 | 1.03 | 0.96 – 1.10 | 0.41 |
| **HIV infection** | - | - | - | 0.95 | 0.66 – 1.35 | 0.76 |

Total CMP counts also correlated directly with proportions of CD4 and CD8 T cells expressing markers of activation, exhaustion and senescence [spearman rho (p value): CD8 activation 0.49 (0.0004), exhaustion 0.39 (0.041), senescence 0.44 (0.0006); CD4 activation 0.44 (0.071), exhaustion 0.27 (0.0007), senescence 0.44 (0.01)]. However, there was no association between total CMPs and monocyte subsets [spearman rho (p value): classical 0.08 (0.57), intermediate −0.018 (0.90), nonclassical −0.07 (0.63)].

### Relationship between CMP subsets, HIV and arterial stiffness

The largest elevations in CMP subsets amongst PLWH were seen with endothelial and platelet derived microparticles. Endothelial derived E-selectin+ CMPs were 1.3-fold higher [median (IQR) 13.0 log particles/mL (12.5 – 14.3) vs 9.9 (8.9 – 10.5); p<0.0001]. The increase in CMPs expressing PECAM but not E-selectin was less significant (Figure 3). Platelet derived CD42a+ CMPs were 1.4-fold higher amongst PLWH [median (IQR) CD42a 15.1 log particles/mL (13.7 – 16.3) vs 10.5 (9.4 – 11.6); p<0.0001]. Amongst leucocyte derived CMPs, CD66b+ and CD16+ CD14- were 1.4-fold and 1.5-fold higher respectively [median (IQR) CD66b: 11.9 log particles/mL (11.3 – 12.7) versus 8.8 (7.9 – 9.3), p<0.0001. CD16+CD14-: 13.1 (11.3 – 133.9) vs 8.7 (7.4 – 9.1), p<0.0001]. For CD14 positive CMPs, only those expressing tissue factor were significantly higher [1.4-fold; median (IQR) 10.1 log particles/mL (8.3 – 11.3) vs 7.0 (6.2 – 8.1), p<0.0001].

**Figure 3.**
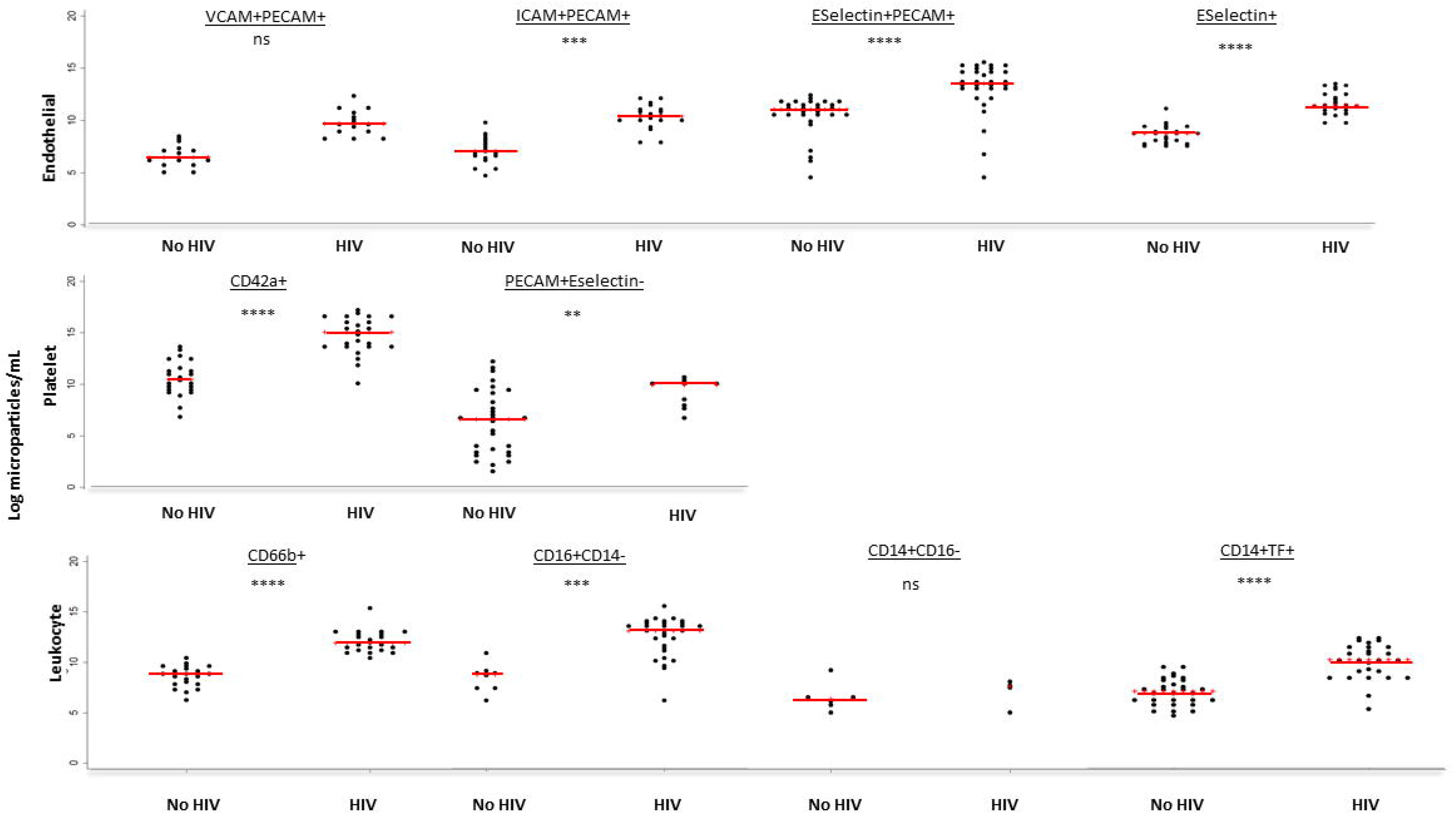
Microparticle subsets according to HIV Status for 69 Malawian Adults. **Figure 3 Legend:** PLWH=People living with HIV. No HIV= People living without HIV. Red lines represent median microparticles count. Grey Asterix represent p value: ns denotes p>0.003, * denotes p<0.003 but >0.0001, **denotes p<0.0001, ***denotes p<0.0001. TF=Tissue Factor. CD=Cluster of Differentiation. VCAM=vascular cell adhesion molecule 1. ICAM=intracellular cell adhesion molecule 1. PECAM=platelet endothelial cell adhesion molecule.

CMPs of endothelial and platelet origin associated closely with cfPWV [spearman rho: E-selectin+ 0.57, p<0.0001 and CD42a 0.56, p<0.0001; Table 3). CD16 positive CD14 negative CMPs also correlated significantly with cfPWV (spearman rho 0.69, p<0.0001).

**Table 3.** Correlations between microparticle subsets and cfPWV.

|  |  | **Total** |  |
| --- | --- | --- | --- |
| **Circulating Microparticle subset** |  | **Spearman's rho** | **P value** |
| **Endothelial** | PECAM+CD42a- | 0.63 | <0.0001 |
|  | Eselectin+ | 0.61 | <0.0001 |
|  | PECAM+Eselectin- | -0.34 | 0.004 |
|  | PECAM+Eselectin+ | 0.55 | <0.0001 |
|  | PECAM-Eselectin+ | 0.53 | <0.0001 |
|  | ICAM+PECAM+ | 0.33 | 0.02 |
|  | ICAM+PECAM- | 0.58 | <0.0001 |
|  | VCAM+PECAM+ | 0.26 | 0.07 |
|  | VCAM+PECAM- | 0.46 | <0.001 |
| **Leucocyte** | CD66b+PECAM- | 0.39 | <0.01 |
|  | CD14-CD16+ | 0.74 | <0.0001 |
|  | CD14+CD16+ | 0.08 | 0.78 |
|  | CD14+CD16- | -0.06 | 0.85 |
| **Platelet** | CD42a+ PECAM- | 0.61 | <0.0001 |

## Discussion

Here we show that total CMPs correlate closely with arterial stiffness and are markedly increased amongst people who present with HIV and advanced immune suppression in Malawi. Microparticles originating from endothelium and platelets were especially high amongst people with HIV and correlated strongly with arterial stiffness; suggesting that these pathways might be prioritised for future studies seeking to reduce inflammation driven cardiovascular risk amongst PLWH during ART initiation.

Each log increase in CMPs showed a 20% increase in cfPWV. A relationship between total CMPs and activated T cells was also demonstrated, in keeping with our previous data suggesting a role for activated and exhausted T cells in increased arterial stiffness amongst PLWH (5). However, the association between CMPs and arterial stiffness was not attenuated by HIV, suggesting that other factors may be at play in the generation of CMPs in this Malawian population.

CMPs are induced in response to cellular stress in the form of activation, apoptosis or physical sheer stress (25). A similar case control study of 15 PLWH established on ART in the USA, showed significantly elevated levels of all types of microparticles compared to 15 participants without HIV infection(23), where microparticles from PLWH directly impaired endothelial cell function. A larger study of PLWH from the Czech Republic also found an increase in microparticles compared to participants without HIV, but found no difference amongst those on ART compared to those who were at their first presentation (26). However, none of these studies assessed the relationship between CMPs and arterial stiffness. By presenting associations with a validated physiological biomarker, we provide evidence for the use of CMPs as a clinically relevant tool to support characterisation of inflammation amongst PLWH.

As well as acting as biomarkers of inflammation, microparticles have been implicated in mediating inflammation induced pathogenesis. They may interact with endothelial cell surface adhesion molecules and cause endothelial damage through the production of nitrous oxide and pro-inflammatory cytokines (27). CMPs have been shown to transfer important material such as miRNA and lipids to other cells (28). In particular, the transfer of the CCR5 receptor to endothelial cells from leucocyte derived microparticles potentially renders them permissible to direct HIV infection (29). Circulating microparticles produced in response to HIV could therefore lead to endothelial damage, and therefore increased risk of cardiovascular disease, through multiple mechanisms including direct endothelial adhesion and activation, transfer of cytotoxic viral proteins, and propagation of low level viral replication.

We discovered that microparticles of endothelial and platelet origin are particularly expanded amongst PLWH in this low income SSA setting. Defining endothelial microparticles as CD51+, da Silva *et al* previously reported endothelial microparticles 20 times higher in PLWH compared to people without HIV (19). Endothelial microparticles promote expression of adhesion molecules on the endothelial cell surface, generating thrombosis, platelet activation and recruitment of inflammatory cells (22). Platelet driven thrombosis in combination with endothelial activation may take advantage of a compromised endothelial barrier and lead to decreased elasticity and smooth muscle reactivity within the arterial wall (30). Inflammatory cytokine release in response to ongoing cellular recruitment, as well as from direct effects of the HIV virus, can also activate and increase the proliferation and migration of vascular smooth muscle cells, and thus induce arterial stiffness (31). Wheway *et al* have previously found endothelial microparticles to form conjugates with T cell subsets, with increased binding to those cells that were pre-stimulated (32). Binding was VCAM and ICAM dependent and endothelial microparticles were able to stimulate in vitro proliferation of T cells.

Although this study examines a small convenience sample, we demonstrate quantification and characterisation of circulating microparticles to be clinically relevant and also confirm that this applies in the low income sub-Saharan Africa setting. We also elucidate the platelet – endothelial axis as an interesting pathway worthy of further investigation in inflammation during early ART. This cohort of people living with HIV had experienced advanced immune suppression and so factors other than the direct effect of HIV itself (e.g. CMV, TB, cryptococcal disease) may be contributing. Further, findings are not generalisable to cohorts with more robust CD4 counts and therefore this needs to be established.

Overall, the characterisation of microparticles in this study lends weight to a model where active and significant immune activation amongst people living with HIV is strongly related to endothelial damage and may involve both CD8 T cell and platelet activation. Further research should investigate whether CMPs might represent translational targets to reduce inflammation driven cardiovascular risk amongst people living with HIV.

## Data Availability

All relevant data is included in the published manuscript or supporting material

## References

1. Farahani M, Mulinder H, Farahani A, Marlink R. Prevalence and distribution of non-AIDS causes of death among HIV-infected individuals receiving antiretroviral therapy: a systematic review and meta-analysis. International journal of STD & AIDS. 2017;28(7):636–50. Epub 2016/02/13.

2. Kuller LH, Tracy R, Belloso W, De Wit S, Drummond F, Lane HC, et al. Inflammatory and coagulation biomarkers and mortality in patients with HIV infection. PLoS Medicine. 2008;5(10):e203. Epub 2008/10/24.

3. Friis-Moller N, Sabin CA, Weber R, d’Arminio Monforte A, El-Sadr WM, Reiss P, et al. Combination antiretroviral therapy and the risk of myocardial infarction. The New England journal of medicine. 2003;349(21):1993–2003. Epub 2003/11/25.

4. Triant VA, Meigs JB, Grinspoon SK. Association of C-reactive protein and HIV infection with acute myocardial infarction. Journal of acquired immune deficiency syndromes (1999). 2009;51(3):268–73. Epub 2009/04/24.

5. Kelly C, Mwandumba HC, Heyderman RS, Jambo K, Kamng’ona R, Chammudzi M, et al. HIV-related arterial stiffness in Malawian adults is associated with proportion of PD-1 expressing CD8 T-cells and reverses with anti-retroviral therapy. J Infect Dis. 2019. Epub 2019/01/11.

6. Craik A, Patel P, Mallewa J, Malisita K, Bitilinyu-Bangoh J, van Oosterhout JJ, et al. Challenges with targeted viral load testing for medical inpatients at Queen Elizabeth Central Hospital in Blantyre, Malawi. Malawi Med J. 2016;28(4):179–81. Epub 2016/01/01.

7. Mugyenyi P, Walker AS, Hakim J, Munderi P, Gibb DM, Kityo C, et al. Routine versus clinically driven laboratory monitoring of HIV antiretroviral therapy in Africa (DART): a randomised non-inferiority trial. Lancet. 2010;375(9709):123–31. Epub 2009/12/17.

8. Heikinheimo T, Chimbayo D, Kumwenda JJ, Kampondeni S, Allain TJ. Stroke outcomes in Malawi, a country with high prevalence of HIV: a prospective follow-up study. PLoS One. 2012;7(3):e33765. Epub 2012/04/06.

9. Benjamin LA, Corbett EL, Connor MD, Mzinganjira H, Kampondeni S, Choko A, et al. HIV, antiretroviral treatment, hypertension, and stroke in Malawian adults: A case-control study. Neurology. 2016;86(4):324–33. Epub 2015/12/20.

10. Laurent S, Alivon M, Beaussier H, Boutouyrie P. Aortic stiffness as a tissue biomarker for predicting future cardiovascular events in asymptomatic hypertensive subjects. Annals of medicine. 2012;44 Suppl 1:S93-7. Epub 2012/06/22.

11. Laurent S, Boutouyrie P, Asmar R, Gautier I, Laloux B, Guize L, et al. Aortic stiffness is an independent predictor of all-cause and cardiovascular mortality in hypertensive patients. Hypertension. 2001;37(5):1236–41. Epub 2001/05/23.

12. Peterson I, Ming D, Kelly C, Malisita K, Mallewa J, Mwandumba HC, et al. Unstructured treatment interruption: an important risk factor for arterial stiffness in adult Malawian patients with antiretroviral treatment. AIDS (London, England). 2016;30(15):2373–8. Epub 2016/07/19.

13. Laurent S, Briet M, Boutouyrie P. Arterial stiffness as surrogate end point: needed clinical trials. Hypertension. 2012;60(2):518–22. Epub 2012/06/27.

14. Laurent S, Cockcroft J, Van Bortel L, Boutouyrie P, Giannattasio C, Hayoz D, et al. Expert consensus document on arterial stiffness: methodological issues and clinical applications. European heart journal. 2006;27(21):2588–605. Epub 2006/09/27.

15. Sima AV, Stancu CS, Simionescu M. Vascular endothelium in atherosclerosis. Cell and tissue research. 2009;335(1):191–203. Epub 2008/09/18.

16. Doran AC, Meller N, McNamara CA. Role of smooth muscle cells in the initiation and early progression of atherosclerosis. Arteriosclerosis, thrombosis, and vascular biology. 2008;28(5):812–9. Epub 2008/02/16.

17. Vion AC, Ramkhelawon B, Loyer X, Chironi G, Devue C, Loirand G, et al. Shear stress regulates endothelial microparticle release. Circulation research. 2013;112(10):1323–33. Epub 2013/03/29.

18. Zwaal RFA, Comfurius P, Bevers EM. Surface exposure of phosphatidylserine in pathological cells. Cellular and Molecular Life Sciences CMLS. 2005;62(9):971–88.

19. da Silva EF, Fonseca FA, Franca CN, Ferreira PR, Izar MC, Salomao R, et al. Imbalance between endothelial progenitors cells and microparticles in HIV-infected patients naive for antiretroviral therapy. AIDS (London, England). 2011;25(13):1595–601. Epub 2011/06/16.

20. Mayne E, Funderburg NT, Sieg SF, Asaad R, Kalinowska M, Rodriguez B, et al. Increased platelet and microparticle activation in HIV infection: upregulation of P-selectin and tissue factor expression. Journal of acquired immune deficiency syndromes (1999). 2012;59(4):340–6. Epub 2011/12/14.

21. Baker JV, Huppler Hullsiek K, Bradford RL, Prosser R, Tracy RP, Key NS. Circulating levels of tissue factor microparticle procoagulant activity are reduced with antiretroviral therapy and are associated with persistent inflammation and coagulation activation among HIV-positive patients. Journal of acquired immune deficiency syndromes (1999). 2013;63(3):367–71. Epub 2013/03/20.

22. Lopez M, San Roman J, Estrada V, Vispo E, Blanco F, Soriano V. Endothelial dysfunction in HIV infection--the role of circulating endothelial cells, microparticles, endothelial progenitor cells and macrophages. AIDS reviews. 2012;14(4):223–30. Epub 2012/12/22.

23. Hijmans JG, Stockelman KA, Garcia V, Levy MV, Brewster LM, Bammert TD, et al. Circulating Microparticles Are Elevated in Treated HIV-1 Infection and Are Deleterious to Endothelial Cell Function. J Am Heart Assoc. 2019;8(4):e011134. Epub 2019/02/20.

24. Van Bortel LM, Laurent S, Boutouyrie P, Chowienczyk P, Cruickshank JK, De Backer T, et al. Expert consensus document on the measurement of aortic stiffness in daily practice using carotid-femoral pulse wave velocity. Journal of hypertension. 2012;30(3):445–8. Epub 2012/01/27.

25. Martin S, Tesse A, Hugel B, Martinez MC, Morel O, Freyssinet JM, et al. Shed membrane particles from T lymphocytes impair endothelial function and regulate endothelial protein expression. Circulation. 2004;109(13):1653–9. Epub 2004/03/17.

26. Snopkova S, Matyskova M, Havlickova K, Jarkovsky J, Svoboda M, Zavrelova J, et al. Increasing procoagulant activity of circulating microparticles in patients living with HIV. Med Mal Infect. 2019. Epub 2019/10/16.

27. Khaddaj Mallat R, Mathew John C, Kendrick DJ, Braun AP. The vascular endothelium: A regulator of arterial tone and interface for the immune system. Crit Rev Clin Lab Sci. 2017;54(7-8):458–70. Epub 2017/11/01.

28. Diehl P, Fricke A, Sander L, Stamm J, Bassler N, Htun N, et al. Microparticles: major transport vehicles for distinct microRNAs in circulation. Cardiovascular research. 2012;93(4):633–44. Epub 2012/01/20.

29. Mack M, Kleinschmidt A, Bruhl H, Klier C, Nelson PJ, Cihak J, et al. Transfer of the chemokine receptor CCR5 between cells by membrane-derived microparticles: a mechanism for cellular human immunodeficiency virus 1 infection. Nat Med. 2000;6(7):769–75. Epub 2000/07/11.

30. Brown RA, Shantsila E, Varma C, Lip GY. Epidemiology and pathogenesis of diffuse obstructive coronary artery disease: the role of arterial stiffness, shear stress, monocyte subsets and circulating microparticles. Annals of medicine. 2016;48(6):444–55. Epub 2016/06/11.

31. Mazzuca P, Caruso A, Caccuri F. HIV-1 infection, microenvironment and endothelial cell dysfunction. New Microbiol. 2016;39(3):163–73. Epub 2016/10/06.

32. Wheway J, Latham SL, Combes V, Grau GE. Endothelial microparticles interact with and support the proliferation of T cells. J Immunol. 2014;193(7):3378–87. Epub 2014/09/05.

